# Childhood Hepatitis B Immunization Coverage and Barriers in Sierra Leone, Liberia, and Guinea: Analysis of National Surveys (2018-2020)

**DOI:** 10.1101/2023.03.16.23287374

**Authors:** George A. Yendewa, Peter B. James, Amir M. Mohareb, Umaru Barrie, Samuel P.E. Massaquoi, Sahr A. Yendewa, Manal Ghazzawi, Tahir Bockarie, Peterlyn E. Cumming, Ibrahima S. Diallo, Ambulai Johnson, Benjamin Vohnm, Lawrence S. Babawo, Gibrilla F. Deen, Mustapha Kabba, Foday Sahr, Sulaiman Lakoh, Robert A. Salata

**Affiliations:** Department of Medicine, Case Western Reserve University School of Medicine, Cleveland, Ohio, USA; Division of Infectious Diseases and HIV Medicine, University Hospitals Cleveland Medical Center, Cleveland, Ohio, USA; Johns Hopkins Bloomberg School of Public Health, Baltimore, Maryland, USA; Faculty of Health, Southern Cross University, Lismore, Australia; Center for Global Health, Massachusetts General Hospital. Boston, Massachusetts, USA; Division of Infectious Diseases, Massachusetts General Hospital, Boston, Massachusetts, USA; Department of Medicine, Harvard Medical School, Boston, Massachusetts, USA; University of Texas Southwestern Medical Center, Dallas, Texas, USA; Ministry of Health and Sanitation, Freetown, Sierra Leone; KnowHep Foundation, Freetown, Sierra Leone; Institut de Nutrition et Santé de L’Enfant (INSE), Conakry, Guinea; National Public Health Institute, Monrovia, Liberia; Department of Nursing, School of Community Health Sciences, Njala University, Bo Campus, Sierra Leone; Connaught Hospital, University of Sierra Leone Teaching Hospitals Complex, Ministry of Health and Sanitation, Freetown, Sierra Leone; Department of Medicine, College of Medicine and Allied Health Sciences, University of Sierra Leone, Freetown, Sierra Leone

**Keywords:** Hepatitis B virus, immunization, Sierra Leone

## Abstract

Vaccination against hepatitis B virus (HBV) is effective at preventing mother-to-child transmission. Sierra Leone, Liberia and Guinea are hyperendemic West African countries; yet childhood immunization coverage is suboptimal and barriers to immunization are poorly understood. We analyzed national survey data (2018-2020) of children aged 4-35 months to assess full HBV immunization (receiving 3 doses of the pentavalent vaccine) and incomplete immunization (receiving < 3 doses). Statistical analysis was conducted using the complex sample command in SPSS (version 28). Multivariate logistic regression was used to identify determinants of incomplete immunization. Overall, 11181 mothers were analyzed (4846 from Sierra Leone, 2788 from Liberia and 3547 from Guinea). Sierra Leone had the highest HBV childhood immunization coverage (70.3%), followed by Liberia (64.5%) and Guinea (40.0%). Within countries, immunization coverage varied by sociodemographic characteristics (education, religion, household wealth index, access to mass media) and healthcare access (antenatal visits, place of delivery and health facility proximity). In multivariate regression analysis, Muslim mothers, lower household wealth index, < 4 antenatal visits, home delivery and distance to health facility were predictors of incomplete immunization (all p < 0.05). Addressing these socioeconomic and healthcare access barriers will be essential to help achieve the 2030 viral hepatitis elimination goals.

## INTRODUCTION

The global burden of hepatitis B virus (HBV) remains a major public health and developmental challenge. According to World Health Organization (WHO) estimates, there were 296 million chronic cases of HBV worldwide in 2021 [1]. Sub-Saharan Africa (SSA) is disproportionately impacted by the HBV epidemic—about 6.1% of the population live with chronic HBV infection, which accounts for approximately 20% of the global burden[1]. In SSA, HBV infection is most commonly acquired vertically through mother-to-child-transmission (MTCT) or horizontally through close contact during early childhood [1, 2]. In response to the global problem of HBV, the Global Health Sector Strategy [3] and the Sustainable Development Goals [4] have endorsed the elimination of HBV as a public health threat by the year 2030.

The most effective method of preventing HBV is through vaccination [4]. The timely administration of the HBV birth dose vaccine given within the first 24 hours of life followed by an additional 2 or 3 doses is 98%-100% effective at preventing MTCT and has been instrumental in reducing the incidence of HBV infection globally [1, 5]. The WHO and other international guidelines recommend universal HBV screening of pregnant women and vaccination of all non-immune newborns and adults [6-8]. However, in many countries in SSA, the HBV vaccine is administered as part of the pentavalent vaccine along with diphtheria, tetanus, pertussis, and Hemophilus influenza type B vaccines at 6, 10, and 14 weeks after birth [9]. However, despite the availability of the HBV vaccine for four decades, immunization coverage in many countries in SSA remains low. Several factors have contributed to the low coverage rates, including inadequate immunization program funding, fragile health systems, and limited knowledge of the risks of HBV infection and the benefits of immunization [9-11]. Understanding and addressing these barriers will be crucial to meeting the HBV elimination goals.

Sierra Leone, Liberia and Guinea are neighboring West African countries with similar cultural, socio-economic and demographic characteristics. Guinea is the most populous of the three countries (15.5 million), while Sierra Leone and Liberia had population estimates of 8.14 million and 5.18 million in 2021, respectively [12]. Furthermore, the three countries share a recent history of civil warfare (1990-2001) and new public health challenges, including the largest Ebola outbreak in history (2014-2016)—all of which have aligned to further weaken already fragile local healthcare systems [13, 14]. Additionally, all three countries are grappling with major HBV epidemics. According to recent estimates, the national prevalence of chronic HBV cases was 13.0% in Sierra Leone [15], 13.5% in Liberia [16] and 11.7% in Guinea [16], indicating hyperendemic levels of HBV infection in all three countries. Guinea initiated childhood HBV vaccination in 2006, followed by Sierra Leone in 2007 and Liberia in 2008 [9]; however, the HBV birth dose vaccine has not been introduced into the childhood immunization schedule and all three countries are yet to implement national HBV control policies due to limited resources and other constraints [9].

Prevention of MTCT of HBV through HBV vaccination is essential to achieve the viral hepatitis elimination goals in Sierra Leone, Guinea and Liberia. However, there is a paucity of studies assessing HBV immunization coverage rates and barriers to the uptake of HBV vaccination in the population. To better inform immunization strategies in Sierra Leone, Liberia and Guinea, we used recent national survey data from the three countries to estimate national HBV immunization coverage levels. Furthermore, we explored sociodemographic, economic and healthcare factors serving as barriers to HBV immunization uptake in the three endemic West African countries.

## METHODS

### Conceptual Framework

Our conceptual framework was guided by the *Strategic framework for research on immunization in the WHO African Region* - *Immunization and Vaccine Development* by the WHO [17] and other relevant literature that have described facilitators and barriers to childhood immunization in low- and middle-income countries [18]. These frameworks outline crucial objectives, such as enhancing vaccine safety and efficacy, promoting research and development of new vaccines, strengthening immunization programs, improving vaccine supply and delivery, and increasing community engagement and demand for immunization services [17, 18]. Facilitators and barriers that have been strongly associated with immunization coverage include lack of knowledge and awareness among the population, vaccine hesitancy, inadequate health infrastructure and human resources, social and cultural factors, and limited access to vaccines and immunization services [17, 18]. For the purpose of our study, our modified conceptual framework proposes that childhood HBV immunization uptake in Sierra Leone, Liberia and Guinea is determined by sociodemographic and economic factors either directly and/or through the mediating influence of other factors such as healthcare access, health behaviors and access to information (Figure 1). Understanding these factors is the first essential step towards crafting evidence-based policy aimed at improving immunization coverage and ultimately lessening the burden of HBV and other vaccine-preventable diseases in this setting.

**Figure 1.**
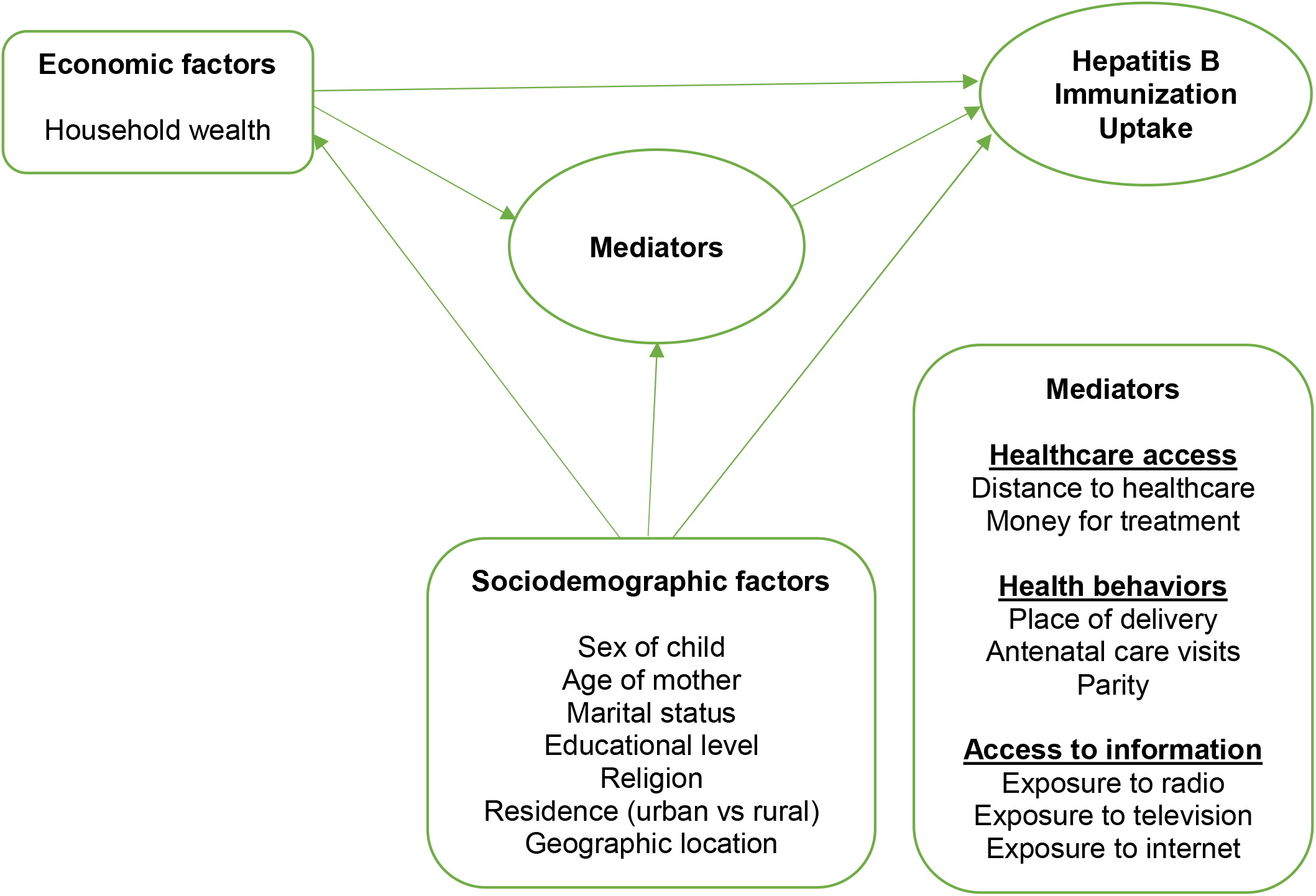
Conceptual framework of complete HBV immunization.

### Survey methodology

We conducted a secondary analysis of data from the most recent Demographic and Health Surveys (DHS) carried out in Sierra Leone (2019), Liberia (2019-2020) and Guinea (2018). The DHS are nationally representative cross-sectional surveys that are regularly conducted in over 90 developing countries to provide information on key population and health indicators. More information on sampling methods and data collection procedures are described in detail in the individual country DHS reports [19-21]. Briefly, the surveys employed a stratified, two-stage cluster probability sampling design and utilized the latest country census sampling frame to identify strata. To ensure representativeness, the size of strata were allocated non-probabilistically, with larger strata “under-sampled” while smaller strata were “over-sampled”. Within each stratum, census enumeration areas were then selected as the primary sampling units in proportion to size. Finally, a fixed number of households were selected within enumeration areas using equal probability systematic sampling. All women aged 15–49 years who permanently resided at or slept in a selected household on the night prior to the survey were eligible to participate. Consenting participants were interviewed using a standardized women’s questionnaire. The interview response rates were > 98% in all three countries [19-21].

### Measures

The primary outcome variables were “complete hepatitis B immunization coverage” and “barriers to incomplete hepatitis B immunization coverage” among children aged 4-35 months in Sierra Leone, Liberia and Guinea, respectively. During the DHS, information on routine childhood immunization was collected by asking eligible mothers about the immunization status of their child (the index case) in the three years preceding the survey. Information on immunization was provided in two ways: (1) written vaccination records or health cards, and (2) verbal reports. The responses provided were recorded as follows: “no”, “vaccination date on card”, “vaccination marked on card”, “reported by mother”, and “don’t know”. To estimate hepatitis B immunization coverage for each of the 3 recommended doses, we assessed the responses recorded under the variables “received pentavalent 1”, “received pentavalent 2”, and “received pentavalent 3”, respectively, given that hepatitis B vaccination is administered as the pentavalent vaccine in Sierra Leone, Liberia and Guinea. The overall coverage for each of the 3 recommended doses was estimated by summing all 3 affirmative responses, i.e., “vaccination date on card”, “vaccination marked on card”, and “reported by mother”. Thus, the primary outcome “complete hepatitis B immunization” was defined as having received all 3 doses of the pentavalent vaccine as reported under the variable “. “Incomplete hepatitis B immunization” was defined as having missed at least 1 of the 3 recommended doses and was a composite of the responses “no” and “don’t know” recorded under the variable “received pentavalent 3”. We excluded children aged < 4 months from the analysis as the third dose of the pentavalent vaccine is administered at 14 weeks.

Where necessary, we recoded variables. The explanatory variables for incomplete HBV vaccination included sex of child (male vs female), mother’s relationship status (recoded as “single” vs “married”), religion (recoded “Christian” vs “Muslim”), educational level (recoded as “no education” vs “primary or higher”), type of place of residence (urban vs rural) and country region. The effect of adolescent motherhood on immunization coverage was assessed as a binary variable (recoded as “15-19 years” vs ≥ “20 years”). Household wealth index was recoded as a binary variable “poor” (poorest and poorer quintiles) vs “rich” (middle, richer and richest quintiles). The number of antenatal visits was recoded as a binary variable (“0-3” vs “≥4”) in accordance with the WHO recommendation of a minimum of 4 antenatal visits for favorable pregnancy outcomes [22]. Parity was similarly recoded as a binary variable (“1-4” vs “5”) in accordance with current regional fertility rates reported by the DHS from each country [19-21]. We assessed the effect of healthcare access on immunization coverage using the variables “money for treatment” (“big problem” vs “not a big problem”) and “distance to health facility” (“big problem” vs “not a big problem”). We recode the variable “place of delivery” as “home” vs “health facility”. Finally, we assessed access to mass media as a source of health information on immunization coverage using the variables “exposure to radio”, “exposure to television” and “exposure to internet” (recoded as “yes” vs “no”, respectively).

### Statistical analyses

We used the complex command package of SPSS Version 28.0 (IBM Corp, NY, USA) to perform statistical analyses, taking into account the complex two-stage cluster sampling design and weighting. Sociodemographic and health characteristics were reported using summary statistics. Categorical variables were expressed as unweighted frequencies (weighted percentages), and vaccination coverage estimates were presented as percentages with 95% confidence intervals (CI). We used Pearson chi-square test to evaluate differences between characteristics. To identify factors associated with incomplete hepatitis B immunization, we employed logistic regression. In the final multivariable logistic regression model, only explanatory variables with p < 0.20 in the univariate analysis were included, with statistical significance set at p < 0.05 for all analyses and presented crude odds ratios (OR) and adjusted odds ratios (aOR) with 95% CI. Furthermore, we assessed statistical collinearity using variance inflation factors and found no evidence of collinearity among explanatory variables (Supplementary file).

### Ethical approval

The protocols for the DHS methodology and data collection procedures were approved by the institutional review boards or ethics committees at ICF Macro, the Sierra Leone Ethics and Scientific Review Committee, the University of Liberia Pacific Institute for Research and Evaluation, and the Guinea National Ethics Committee for Health Research (Comité National d’Ethique pour la Recherche en Santé). Written permission to use the respective country datasets was granted by the DHS.

## RESULTS

### Characteristics of survey respondents

Of a total of 11,181 mothers were included in the analysis, 4846 were from Sierra Leone, 2788 from Liberia and 3547 were from Guinea (Table 1). The children were aged 4-35 months, and the ratio of males to females was roughly 1:1 across countries. A minority of respondents (9.4%-11.7%) were adolescent mothers and single (6.6%-33.5%). Guinea had the highest proportion of mothers with no education (73.2%), compared with Sierra Leone (51.6%) and Liberia (33.5%) (p <0.001). The majority of respondents were Muslim in Sierra Leone (79.6%) and Guinea (87.9%), whereas in Liberia, Christians formed the majority (82.5%). Roughly half (52.9%) of households selected in Liberia were in urban areas, whereas in Sierra Leone and Guinea about two-thirds of households were in rural areas. In each country, an equal proportion of households were selected across wealth quintiles.

**Table 1.** Baseline sociodemographic and health characteristics of index child and households by country

| Characteristics | Sierra Leone | Liberia | Guinea | p-Values |
| --- | --- | --- | --- | --- |
| **Unweighted N (weighted %)** | 4846 (100) | 2788 (100) | 3547 (100) |  |
| **Sex of child** |  |  |  |  |
| Male | 2441 (50.0) | 1358 (49.3) | 1843 (51.7) | 0.110 |
| Female | 2405 (50.0) | 1430 (50.7) | 1708 (48.3) |  |
| **Age of mother, years** |  |  |  |  |
| 15-19 | 455 (9.4) | 346 (11.7) | 378 (11.0) | <0.001 |
| 20-34 | 3310 (69.1) | 1768 (66.7) | 2374 (67.0) |  |
| ≥ 35 | 1081 (21.5) | 674 (21.6) | 795 (22.0) |  |
| **Mother's current marital status** |  |  |  |  |
| Single | 747 (15.5) | 841 (33.5) | 231 (6.6) | <0.001 |
| Married | 4099 (84.5) | 1947 (66.5) | 3316 (93.4) |  |
| **Mother's educational level** |  |  |  |  |
| No education | 2625 (51.6) | 1108 (33.5) | 2592 (73.2) | <0.001 |
| Primary | 707 (15.3) | 877 (26.8) | 432 (11.6) |  |
| Secondary | 1384 (30.3) | 762 (36.3) | 436 (12.6) |  |
| Higher | 130 (2.8) | 41 (3.4) | 87 (2.7) |  |
| **Mother's religion** |  |  |  |  |
| Christian | 967 (20.4) | 2353 (82.5) | 305 (10.5) | <0.001 |
| Muslim | 3877 (79.6) | 364 (14.9) | 3207 (87.9) |  |
| Other | 2 (0) | 71 (2.6) | 35 (1.6) |  |
| **Household wealth index (quintile)** |  |  |  |  |
| Poorest | 1209 (22.8) | 917 (24.0) | 819 (21.8) | <0.001 |
| Poorer | 1119 (21.7) | 773 (21.9) | 760 (21.9) |  |
| Middle | 1021 (20.5) | 553 (18.1) | 651 (18.8) |  |
| Richer | 876 (18.9) | 315 (18.9) | 720 (19.8) |  |
| Richest | 621 (16.2) | 230 (17.0) | 597 (17.6) |  |
| **Type of residence** |  |  |  |  |
| Urban | 1497 (35.3) | 932 (52.9) | 1096 (31.2) | <0.001 |
| Rural | 3349 (64.7) | 1856 (47.1) | 2451 (68.8) |  |
| **Number of antenatal visits** |  |  |  |  |
| 0-3 | 390 (8.4) | 334 (9.8) | 1963 (54.5) |  |
| 4+ | 3605 (74.1) | 2183 (81.4) | 1171 (33.7) |  |
| Don't know/missing data | 851 (17.6) | 271 (8.7) | 413 (11.9) |  |
| **Parity** |  |  |  |  |
| 1-4 | 3593 (74.7) | 1876 (72.9) | 2488 (70.5) | <0.001 |
| 5+ | 1253 (25.3) | 912 (27.1) | 1059 (29.5) |  |
| **Place of delivery** |  |  |  |  |
| Home | 702 (14.0) | 470 (16.3) | 1631 (43.6) | <0.001 |
| Government hospital | 1367 (28.8) | 673 (26.7) | 420 (11.2) |  |
| Government health center | 2126 (44.1) | 219 (5.5) | 915 (27.0) |  |
| Government post | 531 (10.4) | 2 (0.1) | 377 (12.4) |  |
| Private facility | 120 (2.7) | 1424 (51.5) | 204 (5.8) |  |
| **Money for treatment** |  |  |  |  |
| Big problem | 3533 (71.3) | 1190 (38.3) | 2213 (62.0) | <0.001 |
| Not a big problem | 1313 (28.7) | 1598 (61.7) | 1334 (38.0) |  |
| **Distance to health facility** |  |  |  |  |
| Big problem | 2433 (48.3) | 1077 (32.2) | 1732 (47.5) | <0.001 |
| Not a big problem | 2413 (51.7) | 1711 (67.8) | 1815 (52.5) |  |
| **Exposure to radio** |  |  |  |  |
| Yes | 1871 (40.8) | 1484 (54.4) | 2118 (59.7) | <0.001 |
| No | 2975 (59.2) | 1304 (45.6) | 1429 (40.3) |  |
| **Exposure to television** |  |  |  |  |
| Yes | 953 (22.5) | 697 (30.9) | 1432 (41.6) | <0.001 |
| No | 3893 (77.5) | 2091 (69.1) | 2115 (58.4) |  |
| **Exposure to internet** |  |  |  |  |
| Yes | 383 (8.7) | 329 (19.5) | 448 (12.5) |  |
| No | 4463 (91.3) | 2459 (80.5) | 3099 (87.5) |  |
| **Received first dose of HepB vaccine (pentavalent 1)** |  |  |  |  |
| Vaccination date on card | 3374 (69.2) | 1636 (57.1) | 1446 (42.0) | <0.001 |
| Vaccination marked on card | 81 (1.8) | 85 (3.1) | 76 (2.3) |  |
| Reported by mother | 1095 (22.9) | 779 (29.7) | 638 (18.0) |  |
| No or don't know | 296 (6.1) | 288 (10.1) | 1387 (37.7) |  |
| **Received second dose of HepB vaccine (pentavalent 2)** |  |  |  |  |
| Vaccination date on card | 3097 (63.2) | 1505 (52.6) | 1230 (36.0) | <0.001 |
| Vaccination marked on card | 82 (1.9) | 81 (3.1) | 81 (2.3) |  |
| Reported by mother | 791 (16.6) | 608 (23.1) | 375 (11.4) |  |
| No or don't know | 876 (18.3) | 594 (21.1) | 1861 (50.3) |  |
| **Received third dose of HepB vaccine (pentavalent 3)** |  |  |  |  |
| Vaccination date on card | 2778 (56.9) | 1338 (46.9) | 1036 (30.7) | <0.001 |
| Vaccination marked on card | 81 (1.8) | 72 (2.8) | 89 (2.6) |  |
| Reported by mother | 533 (11.6) | 379 (14.9) | 208 (6.7) |  |
| No or don't know | 1454 (29.7) | 999 (35.4) | 2214 (60.0) |  |
| **Country regions** |  |  |  |  |
| **Sierra Leone** |  |  |  |  |
| Eastern | 963 (21.3) |  | - |  |
| Northern | 1119 (19.6) |  | - |  |
| North Western | 908 (19.0) |  | - |  |
| Southern | 1239 (21.5) |  | - |  |
| Western | 617 (18.6) |  | - |  |
| **Liberia** |  |  |  |  |
| North Western | - | 419 (8.5) | - |  |
| South Central | - | 704 (43.5) | - |  |
| South Eastern A | - | 434 (6.7) | - |  |
| South Eastern B | - | 478 (5.5) | - |  |
| North Central | - | 753 (35.7) | - |  |
| **Guinea** |  |  |  |  |
| Boké | - | - | 490 (10.2) |  |
| Conakry | - | - | 355 (13.1) |  |
| Faranah | - | - | 469 (10.4) |  |
| Kankan | - | - | 554 (17.7) |  |
| Kindia | - | - | 470 (14.8) |  |
| Labé | - | - | 455 (11.9) |  |
| Manou | - | - | 323 (7.0) |  |
| N'Zérékoré | - | - | 431 (15.0) |  |

Regarding health-seeking and healthcare access, mothers in Sierra Leone and Liberia had better health indicators in comparison to Guinea. More mothers in Sierra Leone (74.1%) and Liberia (81.4%) accessed antenatal care services 4 or more times during pregnancy, while in Guinea antenatal care attendance was low (33.7%). Similarly, a higher proportion of mothers from Guinea reported delivery at home (43.6%), compared to 14.0% and 16.3% in Sierra Leone and Liberia, respectively. A larger proportion of respondents from Sierra Leone considered money for healthcare payments and distance to healthcare facilities a greater problem compared to the other two countries. On the other hand, information through mass media (radio, television) was more accessible in Guinea, while internet access was more widely available in Liberia.

### Complete hepatitis B immunization coverage rates by respondent characteristics

Table 2 shows complete hepatitis B immunization coverage by respondent sociodemographic and health characteristics across countries. Sierra Leone had the highest overall immunization coverage at 70.3% (95% CI 68.5-72). In Sierra Leone, complete coverage was significantly higher among older mothers (72.2%, 95% CI 69.5-75.0), Christians (74.5%, 95% CI 71.5-77.0), delivery at private facilities (74.5%, 95% CI 66.8-82.2) and close proximity to a healthcare facility (71.6%, 95% CI 69.9-73.4). The regional variations in immunization coverage were not statistically significant.

**Table 2.** Complete HBV immunization coverage rates overall and disaggregated by sociodemographic and health characteristics

| Characteristics | Sierra Leone |  |  | Liberia |  |  | Guinea |  |  |
| --- | --- | --- | --- | --- | --- | --- | --- | --- | --- |
|  | Coverage*<br>% | 95% CI | p-Value | Coverage*<br>% | 95% CI | p-Value | Coverage*<br>% | 95% CI | p-Value |
| **Overall weighted %** | 70.3 | 68.5-72.1 |  | 64.6 | 61.3-68.0 |  | 39.3 | 36.3-42.4 |  |
| **Sex of child** |  |  |  |  |  |  |  |  |  |
| Male | 68.5 | 66.6-70.3 | 0.053 | 66.9 | 64.3-69.5 | **0.042** | 39.5 | 37.2-41.7 | 0.865 |
| Female | 71.4 | 69.5-73.2 |  | 62.4 | 59.7-65.0 |  | 39.2 | 36.9-41.5 |  |
| **Age of mother, years** |  |  |  |  |  |  |  |  |  |
| 15-19 | 65.2 | 60.8-69.7 | **0.042** | 65.7 | 60.3-71.1 | 0.419 | 41.4 | 36.5-46.3 | 0.104 |
| 20-34 | 69.8 | 68.3-71.4 |  | 63.5 | 61.2-65.8 |  | 40.2 | 38.2-42.2 |  |
| 35-49 | 72.2 | 69.5-75.0 |  | 67.4 | 63.5-71.4 |  | 35.6 | 32.2-38.9 |  |
| **Mother's marital status** |  |  |  |  |  |  |  |  |  |
| Single | 69.3 | 66.0-72.7 | 0.746 | 65.2 | 62.0-68.4 | 0.744 | 44.3 | 37.9-50.8 | 0.174 |
| Married | 70.0 | 68.6-71.4 |  | 64.3 | 62.0-66.6 |  | 39.0 | 37.3-40.6 |  |
| **Mother's educational level** |  |  |  |  |  |  |  |  |  |
| No education | 68.3 | 66.5-70.2 | 0.191 | 60.1 | 56.8-63.4 | 0.161 | 35.4 | 33.5-37.2 | **<0.001** |
| Primary | 72.0 | 68.7-75.3 |  | 64.8 | 61.2-68.4 |  | 39.5 | 34.8-44.3 |  |
| Secondary | 71.2 | 68.8-73.5 |  | 68.4 | 65.3-71.4 |  | 55.6 | 51.0-60.3 |  |
| Higher | 73.8 | 66.3-76.5 |  | 68.1 | 58.1-78.1 |  | 70.7 | 61.3-80.1 |  |
| **Mother's religion** |  |  |  |  |  |  |  |  |  |
| Christian | 74.3 | 71.5-77.0 | **0.012** | 70.9 | 66.3-75.5 | 0.067 | 53.9 | 48.8-59.0 | **<0.001** |
| Muslim | 68.8 | 67.3-70.3 |  | 63.7 | 61.6-65.7 |  | 36.8 | 35.1-38.5 |  |
| **Household wealth index (quintile)** |  |  |  |  |  |  |  |  |  |
| Poorest | 67.0 | 64.2-69.8 | 0.173 | 55.0 | 51.1-59.0 | **0.014** | 23.4 | 20.4-26.4 | **0.001** |
| Poorer | 69.2 | 66.4-72.0 |  | 64.6 | 60.6-68.6 |  | 36.1 | 32.7-39.5 |  |
| Middle | 72.4 | 69.6-75.2 |  | 67.6 | 63.3-71.9 |  | 38.2 | 34.5-41.9 |  |
| Richer | 72.3 | 69.3-75.2 |  | 70.1 | 66.6-74.8 |  | 44.7 | 41.0-48.4 |  |
| Richest | 69.1 | 65.8-72.3 |  | 68.2 | 63.8-72.6 |  | 58.2 | 54.3-62.1 |  |
| **Type of residence** |  |  |  |  |  |  |  |  |  |
| Urban | 70.8 | 68.6-73.0 | 0.493 | 67.6 | 65.1-70.1 | 0.062 | 52.6 | 49.6-55.55 | **<0.001** |
| Rural | 69.4 | 67.8-71.1 |  | 61.3 | 58.5-64.0 |  | 33.3 | 31.4-35.2 |  |
| **Number of antenatal visits** |  |  |  |  |  |  |  |  |  |
| 0-3 | 64.9 | 60.2-69.6 | 0.058 | 47.5 | 41.2-53.7 | **<0.001** | 29.4 | 27.4-31.5 | **0.001** |
| 4+ | 71.0 | 69.5-72.5 |  | 67.4 | 65.3-69.4 |  | 55.3 | 52.4-58.1 |  |
| **Parity** |  |  |  |  |  |  |  |  |  |
| 1-4 | 70.1 | 68.6-71.6 | 0.729 | 66.4 | 64.2-68.6 | **0.049** | 40.9 | 39.0-42.8 | **0.007** |
| 5+ | 69.5 | 66.8-72.1 |  | 59.8 | 56.2-63.5 |  | 35.6 | 32.6-38.5 |  |
| **Place of delivery** |  |  |  |  |  |  |  |  |  |
| Home | 65.9 | 63.3-69.5 | **<0.001** | 51.6 | 46.8-56.4 | **<0.001** | 23.6 | 21.4-25.7 | **0.001** |
| Government hospital | 68.3 | 65.8-70.7 |  | 70.0 | 66.6-73.5 |  | 58.3 | 53.4-63.2 |  |
| Government health center | 71.4 | 69.5-73.3 |  | 72.1 | 64.5-79.7 |  | 49.0 | 45.9-52.2 |  |
| Government post | 72.3 | 68.4-76.3 |  | - | - |  | 52.2 | 47.5-56.9 |  |
| Private facility | 74.5 | 66.8-82.2 |  | 65.1 | 62.5-67.7 |  | 48.3 | 41.4-55.2 |  |
| **Getting medical help for self: money for treatment** |  |  |  |  |  |  |  |  |  |
| Big problem | 69.4 | 67.8-70.9 | 0.112 | 65.3 | 62.3-68.3 | 0.647 | 35.1 | 33.1-37.1 | **<0.001** |
| Not a big problem | 71.2 | 68.8-73.6 |  | 64.2 | 61.8-66.6 |  | 46.2 | 43.6-48.9 |  |
| **Distance to health facility** |  |  |  |  |  |  |  |  |  |
| Big problem | 68.0 | 66.1-70.0 | **0.032** | 64.1 | 60.8-67.4 | 0.773 | 32.1 | 29.9-34.3 | **<0.001** |
| Not a big problem | 71.6 | 69.9-73.4 |  | 64.9 | 62.6-67.1 |  | 45.9 | 43.6-48.1 |  |
| **Exposure to radio** |  |  |  |  |  |  |  |  |  |
| Yes | 70.6 | 68.5-72.6 | 0.514 | 67.3 | 64.8-69.8 | 0.051 | 43.2 | 41.1-45.3 | **<0.001** |
| No | 69.5 | 67.8-71.2 |  | 61.4 | 58.6-64.2 |  | 33.6 | 31.1-36.1 |  |
| **Exposure to television** |  |  |  |  |  |  |  |  |  |
| Yes | 70.3 | 67.5-73.0 | 0.822 | 67.4 | 64.1-70.7 | 0.294 | 47.6 | 45.0-50.2 | **<0.001** |
| No | 69.8 | 68.3-71.3 |  | 63.4 | 61.1-65.6 |  | 33.4 | 31.4-35.5 |  |
| **Exposure to internet** |  |  |  |  |  |  |  |  |  |
| Yes | 69.6 | 65.2-74.1 | 0.918 | 70.8 | 66.8-74.8 | 0.111 | 57.7 | 53.1-62.3 | **<0.001** |
| No | 69.9 | 68.6-71.3 |  | 63.1 | 61.0-65.2 |  | 36.7 | 35.0-38.4 |  |
| **Country regions** |  |  |  |  |  |  |  |  |  |
| **Sierra Leone** |  |  |  |  |  |  |  |  |  |
| Eastern | 73.1 | 70.3-75.8 | 0.058 | - | - |  | - | - |  |
| Northern | 66.5 | 63.4-69.5 |  | - | - |  | - | - |  |
| North Western | 67.6 | 64.5-70.6 |  | - | - |  | - | - |  |
| Southern | 73.1 | 70.4-75.8 |  | - | - |  | - | - |  |
| Western | 68.6 | 65.6-71.7 |  | - | - |  | - | - |  |
| **Liberia** |  |  |  |  |  |  |  |  |  |
| North Western | - | - |  | 70.1 | 63.9-76.2 | 0.427 | - | - |  |
| South Central | - | - |  | 63.9 | 61.0-66.7 |  | - | - |  |
| South Eastern A | - | - |  | 57.7 | 50.2-65.2 |  | - | - |  |
| South Eastern B | - | - |  | 65.8 | 57.8-73.7 |  | - | - |  |
| North Central | - | - |  | 65.4 | 62.3-67.1 |  | - | - |  |
| **Guinea** |  |  |  |  |  |  |  |  |  |
| Boké | - | - |  | - | - |  | 27.6 | 23.0-32.3 | **<0.001** |
| Conakry | - | - |  | - | - |  | 55.5 | 50.9-60.1 |  |
| Faranah |  |  |  | - | - |  | 39.4 | 34.4-44.4 |  |
| Kankan | - | - |  | - | - |  | 48.0 | 44.1-51.9 |  |
| Kindia | - | - |  | - | - |  | 38.8 | 34.6-43.0 |  |
| Labé | - | - |  | - | - |  | 17.1 | 13.5-20.7 |  |
| Manou | - | - |  | - | - |  | 25.1 | 19.6-30.5 |  |
| N'Zérékoré | - | - |  | - | - |  | 47.8 | 43.5-52.1 |  |

\*Complete HBV immunization coverage was defined as completing all 3 series of vaccines administered at 6, 10, and 14 weeks after birth and was estimated as a composite of “vaccination date on card”, “vaccination marked on card” and “reported by mother” (Table 1)

Liberia had an overall hepatitis B immunization coverage rate of 64.5% (95% CI 61.3-68.0). Male children (66.9%,95% CI 64.4-69.5) had higher immunization coverage compared to females (62.4%, 95% CI 59.7-65.0). When disaggregated by respondent characteristics, the highest immunization coverage rates were in households in the richer wealth quintile (70.1%, 95% CI 66.6-74.8), mothers who had ≥ 4 antenatal visits (67.4%, 95% CI 65.2-69.4) and delivery at a government health center (72.1%, 95% CI 64.5-79.7).

Guinea had the lowest overall hepatitis B immunization coverage rate (40.0%, 95% CI 36.9-43.1). Immunization coverage in Guinea was much more heavily influenced by socioeconomic and health factors compared with Sierra Leone and Liberia. The highest immunization coverage was observed among respondents with higher level of education (70.7%, 95% CI 61.3-80.1), Christians (53.9%, 95% CI 48.8-59.0), households in the richest wealth quintiles (58.2%, 95% CI 54.3-62.1), residence in urban areas (52.6%, 95% CI 49.6-55.55), ≥ 4 antenatal visits (55.3%, 95% CI 52.4-58.1), delivery at a government hospital (58.3%, 95% CI 53.4-63.2), access to internet (57.7%, 95% CI 53.1-62.3), mothers from the Conakry region (55.5%, 95% CI 50.9-60.1).

### Predictors of incomplete hepatitis B immunization coverage

Tables 3 and 4 display the results of univariate and multivariate regression analysis of factors associated with incomplete hepatitis B immunization coverage. In Sierra Leone, being an adolescent mother (aOR 1.36, 95% CI 1.05-1.75), Muslim (aOR 1.28, 95% CI 1.03-1.59), lower wealth index (aOR 1.20, 95% CI 1.00-1.43) and distance to health facility (aOR 1.20 95% CI 1.01-1.43) were associated incomplete immunization, while being a female child was a protective factor (aOR 0.86, 95% CI 0.74-0.99). In contrast, female children (aOR 1.83, 95% CI 1.30-2.59), fewer than 4 antenatal visits (aOR 1.83, 95% CI 1.30-2.59) and home delivery (aOR 1.48, 95% CI 1.02-2.13) were significantly associated with incomplete immunization. Similarly in Guinea, healthcare factors were the dominant predictors of incomplete immunization, i.e., < 4 antenatal visits (aOR 2.31, 95% CI 1.87-2.85) and home delivery (aOR 2.18, 95% CI 1.74-2.74). Additionally in Guinea, being Muslim (aOR 1.96, 95% CI 1.28-2.99) and being resident in the Labé region (aOR 2.58, 95% CI 1.49-4.48) predicted incomplete immunization.

**Table 3.**
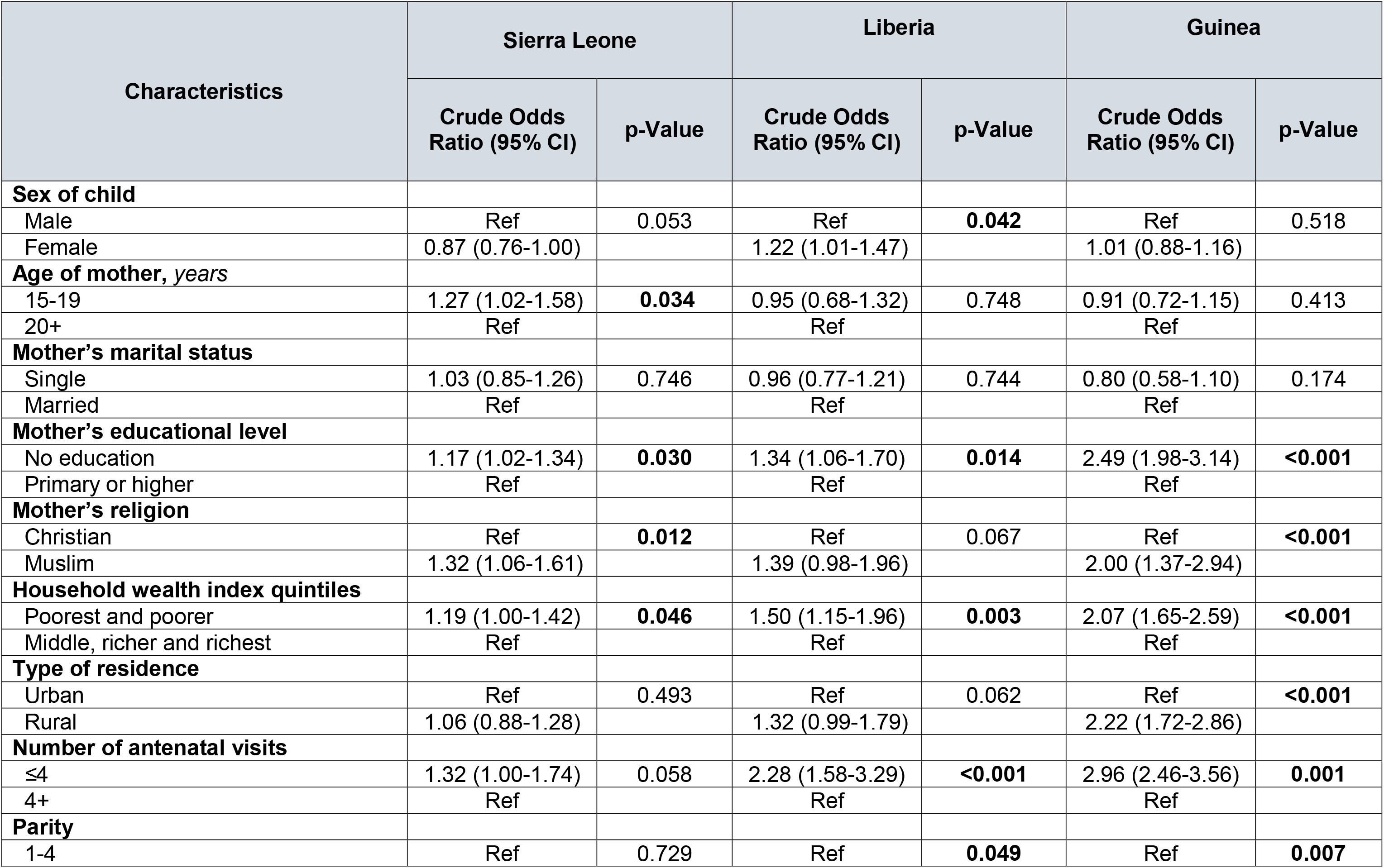
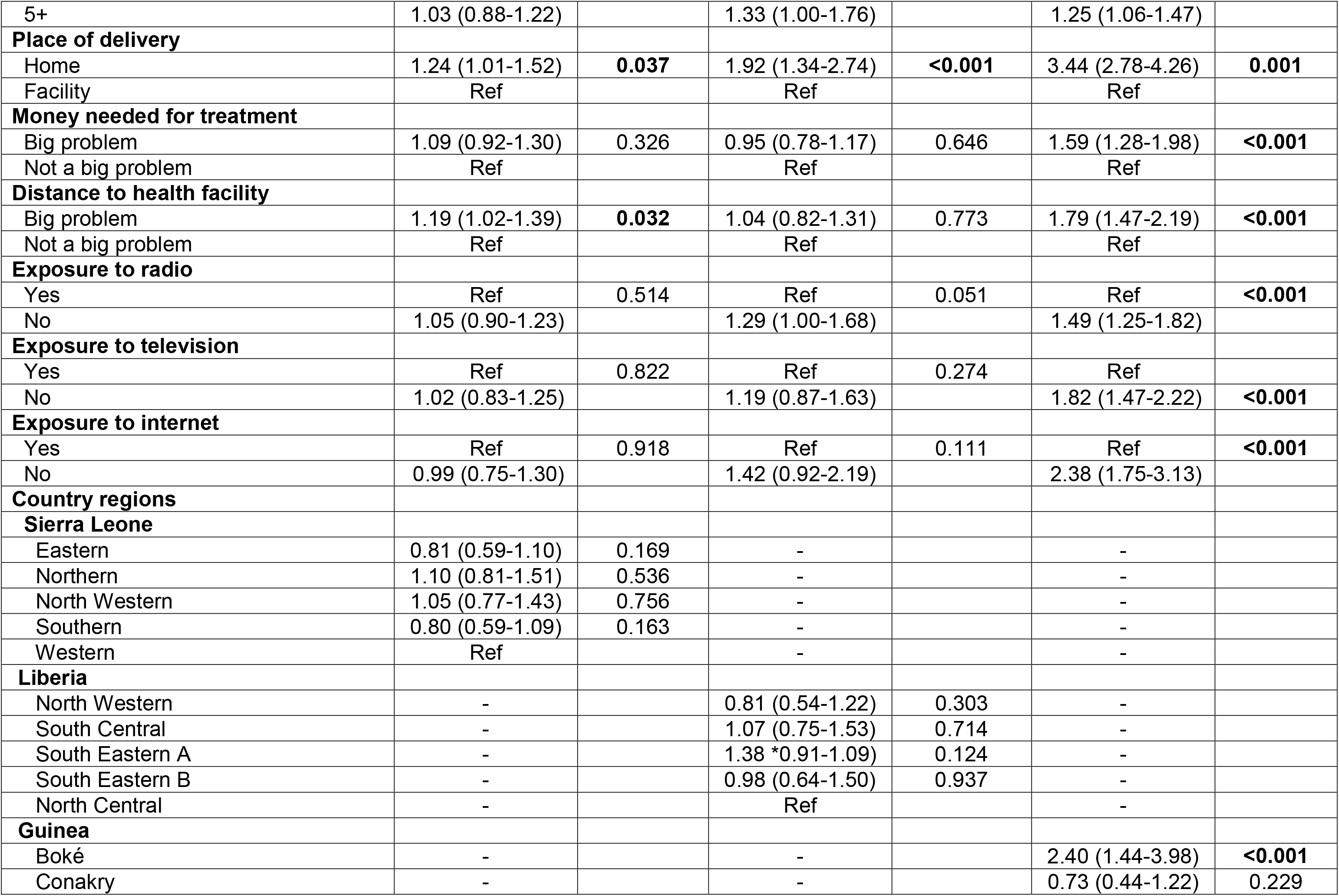
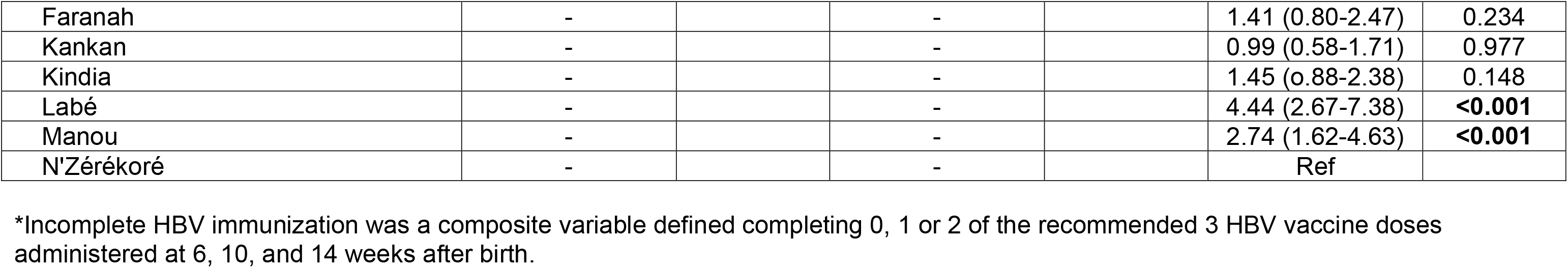
Univariate analysis of factors associated with incomplete HBV immunization

| Characteristics | Sierra Leone |  | Liberia |  | Guinea |  |
| --- | --- | --- | --- | --- | --- | --- |
|  | Crude Odds Ratio (95% CI) | p-Value | Crude Odds Ratio (95% CI) | p-Value | Crude Odds Ratio (95% CI) | p-Value |
| **Sex of child** |  |  |  |  |  |  |
| Male | Ref | 0.053 | Ref | **0.042** | Ref | 0.518 |
| Female | 0.87 (0.76-1.00) |  | 1.22 (1.01-1.47) |  | 1.01 (0.88-1.16) |  |
| **Age of mother, years** |  |  |  |  |  |  |
| 15-19 | 1.27 (1.02-1.58) | **0.034** | 0.95 (0.68-1.32) | 0.748 | 0.91 (0.72-1.15) | 0.413 |
| 20+ | Ref |  | Ref |  | Ref |  |
| **Mother's marital status** |  |  |  |  |  |  |
| Single | 1.03 (0.85-1.26) | 0.746 | 0.96 (0.77-1.21) | 0.744 | 0.80 (0.58-1.10) | 0.174 |
| Married | Ref |  | Ref |  | Ref |  |
| **Mother's educational level** |  |  |  |  |  |  |
| No education | 1.17 (1.02-1.34) | **0.030** | 1.34 (1.06-1.70) | **0.014** | 2.49 (1.98-3.14) | **<0.001** |
| Primary or higher | Ref |  | Ref |  | Ref |  |
| **Mother's religion** |  |  |  |  |  |  |
| Christian | Ref | **0.012** | Ref | 0.067 | Ref | **<0.001** |
| Muslim | 1.32 (1.06-1.61) |  | 1.39 (0.98-1.96) |  | 2.00 (1.37-2.94) |  |
| **Household wealth index quintiles** |  |  |  |  |  |  |
| Poorest and poorer | 1.19 (1.00-1.42) | **0.046** | 1.50 (1.15-1.96) | **0.003** | 2.07 (1.65-2.59) | **<0.001** |
| Middle, richer and richest | Ref |  | Ref |  | Ref |  |
| **Type of residence** |  |  |  |  |  |  |
| Urban | Ref | 0.493 | Ref | 0.062 | Ref | **<0.001** |
| Rural | 1.06 (0.88-1.28) |  | 1.32 (0.99-1.79) |  | 2.22 (1.72-2.86) |  |
| **Number of antenatal visits** |  |  |  |  |  |  |
| ≤4 | 1.32 (1.00-1.74) | 0.058 | 2.28 (1.58-3.29) | **<0.001** | 2.96 (2.46-3.56) | **0.001** |
| 4+ | Ref |  | Ref |  | Ref |  |
| **Parity** |  |  |  |  |  |  |
| 1-4 | Ref | 0.729 | Ref | **0.049** | Ref | **0.007** |
| 5+ | 1.03 (0.88-1.22) |  | 1.33 (1.00-1.76) |  | 1.25 (1.06-1.47) |  |
| **Place of delivery** |  |  |  |  |  |  |
| Home | 1.24 (1.01-1.52) | **0.037** | 1.92 (1.34-2.74) | **<0.001** | 3.44 (2.78-4.26) | **0.001** |
| Facility | Ref |  | Ref |  | Ref |  |
| **Money needed for treatment** |  |  |  |  |  |  |
| Big problem | 1.09 (0.92-1.30) | 0.326 | 0.95 (0.78-1.17) | 0.646 | 1.59 (1.28-1.98) | **<0.001** |
| Not a big problem | Ref |  | Ref |  | Ref |  |
| **Distance to health facility** |  |  |  |  |  |  |
| Big problem | 1.19 (1.02-1.39) | **0.032** | 1.04 (0.82-1.31) | 0.773 | 1.79 (1.47-2.19) | **<0.001** |
| Not a big problem | Ref |  | Ref |  | Ref |  |
| **Exposure to radio** |  |  |  |  |  |  |
| Yes | Ref | 0.514 | Ref | 0.051 | Ref | **<0.001** |
| No | 1.05 (0.90-1.23) |  | 1.29 (1.00-1.68) |  | 1.49 (1.25-1.82) |  |
| **Exposure to television** |  |  |  |  |  |  |
| Yes | Ref | 0.822 | Ref | 0.274 | Ref |  |
| No | 1.02 (0.83-1.25) |  | 1.19 (0.87-1.63) |  | 1.82 (1.47-2.22) | **<0.001** |
| **Exposure to internet** |  |  |  |  |  |  |
| Yes | Ref | 0.918 | Ref | 0.111 | Ref | **<0.001** |
| No | 0.99 (0.75-1.30) |  | 1.42 (0.92-2.19) |  | 2.38 (1.75-3.13) |  |
| **Country regions** |  |  |  |  |  |  |
| **Sierra Leone** |  |  |  |  |  |  |
| Eastern | 0.81 (0.59-1.10) | 0.169 | - |  | - |  |
| Northern | 1.10 (0.81-1.51) | 0.536 | - |  | - |  |
| North Western | 1.05 (0.77-1.43) | 0.756 | - |  | - |  |
| Southern | 0.80 (0.59-1.09) | 0.163 | - |  | - |  |
| Western | Ref |  | - |  | - |  |
| **Liberia** |  |  |  |  |  |  |
| North Western | - |  | 0.81 (0.54-1.22) | 0.303 | - |  |
| South Central | - |  | 1.07 (0.75-1.53) | 0.714 | - |  |
| South Eastern A | - |  | 1.38 *0.91-1.09) | 0.124 | - |  |
| South Eastern B | - |  | 0.98 (0.64-1.50) | 0.937 | - |  |
| North Central | - |  | Ref |  | - |  |
| **Guinea** |  |  |  |  |  |  |
| Boké | - |  | - |  | 2.40 (1.44-3.98) | **<0.001** |
| Conakry | - |  | - |  | 0.73 (0.44-1.22) | 0.229 |
| Faranah | - |  | - |  | 1.41 (0.80-2.47) | 0.234 |
| Kankan | - |  | - |  | 0.99 (0.58-1.71) | 0.977 |
| Kindia | - |  | - |  | 1.45 (0.88-2.38) | 0.148 |
| Labé | - |  | - |  | 4.44 (2.67-7.38) | **<0.001** |
| Manou | - |  | - |  | 2.74 (1.62-4.63) | **<0.001** |
| N'Zérékoré | - |  | - |  | Ref |  |

\*Incomplete HBV immunization was a composite variable defined completing 0, 1 or 2 of the recommended 3 HBV vaccine doses administered at 6, 10, and 14 weeks after birth.

**Table 4.** Multivariate analysis of factors associated with incomplete HBV immunization

| Characteristics | Sierra Leone |  | Liberia |  | Guinea |  |
| --- | --- | --- | --- | --- | --- | --- |
|  | Adjusted Odds Ratio (95% CI) | p-Value | Adjusted Odds Ratio (95% CI) | p-Value | Adjusted Odds Ratio (95% CI) | p-Value |
| **Sex of child** |  |  |  |  |  |  |
| Male | Ref | **0.037** | Ref | **0.041** |  |  |
| Female | 0.86 (0.74-0.99) |  | 1.25 (1.01-1.56) |  |  |  |
| **Age of mother, years** |  |  |  |  |  |  |
| 15-19 | 1.36 (1.05-1.75) | **0.019** |  |  |  |  |
| 20+ | Ref |  |  |  |  |  |
| **Mother's marital status** |  |  |  |  |  |  |
| Single |  |  |  |  | 1.67 (0.85-1.60) | 0.342 |
| Married |  |  |  |  | Ref |  |
| **Mother's educational level** |  |  |  |  |  |  |
| No education | 1.10 (0.94-1.30) | 0.243 | 1.19 (0.91-1.56) | 0.195 | 1.13 (0.87-1.46) | 0.373 |
| Primary or higher | Ref |  | Ref |  | Ref |  |
| **Mother's religion** |  |  |  |  |  |  |
| Christian | Ref | **0.027** | Ref | 0.408 | Ref | **0.002** |
| Muslim | 1.28 (1.03-1.59) |  | 1.18 (0.80-1.72) |  | 1.96 (1.28-2.99) |  |
| **Household wealth index quintiles** |  |  |  |  |  |  |
| Poorest and poorer | 1.20 (1.00-1.43) | **0.050** | 1.27 (0.94-1.71) | 0.126 | 1.20 (0.92-1.58) | 0.179 |
| Middle, richer and richest | Ref |  | Ref |  | Ref |  |
| **Type of residence** |  |  |  |  |  |  |
| Urban |  |  | Ref | 0.671 | Ref | 0.781 |
| Rural |  |  | 1.09 (0.73-1.63) |  | 1.05 (0.73-1.51) |  |
| **Number of antenatal visits** |  |  |  |  |  |  |
| ≤3 | 1.29 (0.98-1.69) | 0.070 | 1.83 (1.30-2.59) | **<0.001** | 2.31 (1.87-2.85) | **<0.001** |
| 4+ | Ref |  | Ref |  | Ref |  |
| **Parity** |  |  |  |  |  |  |
| 1-4 |  |  | 0.85 (0.63-1.16) | 0.303 | 0.97 (0.80-1.16) | 0.707 |
| 5+ |  |  | Ref |  | Ref |  |
| **Place of delivery** |  |  |  |  |  |  |
| Home | 1.08 (0.86-1.36) | 0.527 | 1.48 (1.02-2.13) | **0.038** | 2.18 (1.74-2.74) | **<0.001** |
| Facility | Ref |  | Ref |  | Ref |  |
| **Money needed for treatment** |  |  |  |  |  |  |
| Big problem |  |  |  |  | 1.07 (0.85-1.35) | 0.163 |
| Not a big problem |  |  |  |  | Ref |  |
| **Distance to health facility** |  |  |  |  |  |  |
| Big problem | 1.20 (1.01-1.43) | **0.034** |  |  | 1.17 (0.95-1.44) | 0.574 |
| Not a big problem | Ref |  |  |  | Ref |  |
| **Exposure to radio** |  |  |  |  |  |  |
| Yes |  |  | Ref | 0.289 | Ref | 0.137 |
| No |  |  | 1.17 (0.88-1.55) |  | 1.13 (0.94-1.37) |  |
| **Exposure to television** |  |  |  |  |  |  |
| Yes |  |  |  |  | Ref | 0.245 |
| No |  |  |  |  | 1.16 (0.90-1.49) |  |
| **Exposure to internet** |  |  |  |  |  |  |
| Yes |  |  | Ref | 0.792 | Ref | 0.176 |
| No |  |  | 1.06 (0.67-1.70) |  | 1.27 (0.90-1.79) |  |
| **Country regions** |  |  |  |  |  |  |
| **Sierra Leone** |  |  |  |  |  |  |
| Eastern | 0.84 (0.58-1.21) | 0.340 | - |  | - |  |
| Northern | 1.09 (0.75-1.57) | 0.663 | - |  | - |  |
| North Western | 1.01 (0.70-1.45) | 0.955 | - |  | - |  |
| Southern | 0.75 (0.52-1.09) | 0.133 | - |  | - |  |
| Western | Ref |  | - |  | - |  |
| **Liberia** |  |  |  |  |  |  |
| North Western | - |  | 0.70 (0.47-1.02) | 0.064 | - |  |
| South Central | - |  | 1.22 (0.78-1.89) | 0.737 | - |  |
| South Eastern A | - |  | 1.26 (0.82-1.94) | 0.289 | - |  |
| South Eastern B | - |  | 0.90 (0.57-1.41) | 0.629 | - |  |
| North Central | - |  | Ref |  | - |  |
| **Guinea** |  |  |  |  |  |  |
| Boké | - |  | - |  | 1.38 (0.82-2.32) | 0.228 |
| Conakry | - |  | - |  | 0.79 (0.46-1.37) | 0.402 |
| Faranah | - |  | - |  | 0.72 (0.40-1.29) | 0.263 |
| Kankan | - |  | - |  | 0.65 (0.36-1.18) | 0.158 |
| Kindia | - |  | - |  | 0.96 (0.57-1.61) | 0.871 |
| Labé | - |  | - |  | 2.58 (1.49-4.48) | **<0.001** |
| Manou | - |  | - |  | 1.44 (0.80-2.59) | 0.220 |
| N'Zérékoré | - |  | - |  | Ref |  |

\*Complete HBV immunization coverage was defined as completing all 3 series of vaccines administered at 6, 10, and 14 weeks after birth and was estimated as a composite of “vaccination date on card”, “vaccination marked on card” and “reported by mother”

## DISCUSSION

To the best of our knowledge, this is the first population-based study to estimate national hepatitis B immunization coverage rates and identify barriers to immunization among children in Sierra Leone, Liberia, and Guinea, three West African countries with a high burden of HBV infection. Sierra Leone had the highest national immunization coverage at 70.3%, while Liberia and Guinea had lower coverage rates at 64.7% and 40%, respectively. Our estimates were slightly lower than coverage rates compiled jointly by the WHO and the United Nation’s Children’s Emergency Fund (WHO/UNICEF), which reported a 95% immunization coverage rate for Sierra Leone in 2019, 70% for Liberia in 2020 and 47% for Guinea in 2018 [23]. Furthermore, the coverage rates from our study were lower than those reported by other West African countries such as Senegal (93%), the Gambia (88%), Burkina Faso (91%), and Mali (77%) [23]. The differences in immunization coverage among the three countries could be attributed to various factors, including social-cultural and healthcare service-related factors, as well as variations in the data sources used to calculate the estimates. The WHO/UNICEF estimates relied on both administrative data from healthcare providers and population-based surveys and focused on children aged 12-35 months [23]. In contrast, our estimates were solely based on population-based surveys and included younger children aged 4-35 months, who were more likely to lagging behind in completing the vaccine series [19-21].

Annually, around 820,000 people die from the complications of chronic HBV infection worldwide [1]. In many endemic countries in SSA, there are still entrenched disparities and inequities in access to quality healthcare including HBV screening, treatment and prevention services, despite the availability of an effective HBV vaccine since 1982 and the WHO’s call for the global elimination of viral hepatitis as a public health threat by 2030 [4]. In our examination of disparities in HBV immunization coverage based on key sociodemographic, economic, and healthcare access factors, we identified significant intra- and inter-countries differences in immunization uptake. These factors were most influential within Guinea, and between Guinea and the other countries. Of the factors examined, higher rates of immunization coverage were universally observed for facility-based delivery in all countries, while home deliveries registered the lowest rates. Other factors that influenced higher immunization coverage in more than one country were antenatal care visits, higher household wealth quintiles, fewer births, religion (Christianity), and within close proximity to healthcare facility. In Guinea, higher maternal mother’s level of education, access to information through mass media, and payments for health services were additional factors that determined level of immunization coverage. Our findings are in agreement with studies from other African countries, notably Cameroon, the Central African Republic, Senegal, Nigeria, Ethiopia, and Uganda [24-30].

Furthermore, we assessed predictors of incomplete immunization coverage. Healthcare access indices predominated as were the most important predictors. Children born to mothers who had fewer than four antenatal care visits had 1.83- to 2.3-fold higher odds of not completing immunization in Liberia and Guinea. Similarly, home delivery was significantly associated with incomplete coverage in the two countries. In contrast, in Sierra Leone, children with mothers who considered distance to health facility a “big problem” were significantly less likely to complete the immunization series. These findings are not surprising, as antenatal care attendance and delivery in healthcare facility provides opportunities for educating mothers on the benefits of vaccination and other vital maternal and child health services [31, 32]. Taken all together, our findings are in agreement with others who have suggested that HBV infection is fundamentally a disease of poverty [33, 34]; thus, HBV control efforts should aim to address socioeconomic inequities to achieve elimination targets [9, 33, 34].

A notable finding was the observed impact of religion on HBV immunization uptake. Children of Muslim mothers were less likely to complete the HBV immunization series in Sierra Leone and Guinea, which have majority Muslim populations. Studies from elsewhere corroborate these findings. A recent systematic review from the United Kingdom found that being Muslim played a role in parents’ decision making about routine childhood immunization [35]. However, these findings warrant more careful interpretation, as there is no conclusive evidence to suggest that Muslims are less likely to vaccinate compared with people of other religious or cultural backgrounds. For example, a study from Malaysia found that the majority of Muslim parents surveyed had positive attitudes towards routine childhood immunizations and believed that vaccines were permissible under Islamic law (or “Sharia”) [36]. In contrast, studies have found that Evangelical Christians in the United States [37, 38] and Christians in Nigeria [39] have been the most hesitant to vaccine against coronavirus-19 compared with other religious groups. Overall, these findings suggest that vaccine acceptance is a complex issue that is influenced by a multitude of factors, including education, access to healthcare, trust in healthcare providers, and cultural, religious and political beliefs [35-39]. To improve vaccine uptake among all communities, it is therefore important to address concerns and provide accurate information about vaccine safety and efficacy.

Our study did not directly assess the impact of HBV birth dose vaccination on completion rates as it has not been implemented in any of the three countries examined. However, it is an important factor to consider when examining the suboptimal coverage rates observed in our study population. The WHO reports that as of 2021, only 14 of the 47 countries in the WHO Africa region have integrated the HBV birth dose vaccine into their national immunization programs and about 1 in 5 newborns in this region receive the birth dose vaccine [40]. In addition to preventing MTCT of HBV, multiple studies, mostly from high income settings, have shown that timely administration of the HBV birth dose vaccine is associated with 1.8- to 3-fold higher completion rates of the three-dose HBV vaccine series, as well as other routine childhood vaccines [41-43]. In the few countries in SSA where the HBV birth dose has been implemented, impact assessment studies have highlighted barriers such as implementation cost, a high proportion of births taking place outside of health facilities (e.g., home births), limited access to skilled birth attendants, and inadequate knowledge of HBV among health workers and parents as major factors limiting successful implementation of the birth dose vaccination [40, 43-45]. Thus, effective implementation of HBV birth dose vaccination in SSA requires a comprehensive approach that addresses health system strengthening, awareness-raising, delivery platforms, vaccine supply chain management, and collaboration and partnership. By extension, successful implementation of these strategies can enhance HBV immunization coverage and reduce the burden of HBV infection in SSA.

It is important to acknowledge the limitations of our study, which can impact the generalizability of our findings. These pertain to the use of secondary data sources, potential biases in data collection, challenges in determining causal links, and potential under-sampling of certain populations. To address some of these limitations, it would have been useful to obtain additional sources of survey coverage data and to explore country-specific correlates of complete HBV vaccination in more detail. Nevertheless, the DHS surveys are carefully planned and employ rigorous methodology, which adds to the validity of the study’s findings. Overall, while the limitations should be considered when interpreting the results, our study provides valuable insights into the prevalence and socio-demographic correlates of childhood HBV immunization coverage in the three countries examined.

## CONCLUSIONS

In summary, we found suboptimal levels of HBV immunization among children in Sierra Leone, Liberia and Guinea. Within countries, immunization coverage varied significantly based on sociodemographic characteristics and healthcare access. Being a male child, Muslim mothers, low household wealth index, poor antenatal clinic attendance, home delivery and proximity to health facility were important determinants of incomplete immunization. These findings underscore the critical need to address socioeconomic and healthcare inequities that are driving suboptimal HBV immunization uptake and stalling progress towards achieving the 2030 global viral hepatitis elimination targets in these endemic countries.

## Data Availability

All data produced in the present work are contained in the manuscript

## AUTHOR CONTRIBUTIONS

Conceptualization, G.A.Y., P.B.J., and R.A.S.; methodology, G.A.Y., P.B.J., A.M.M., U.B., S.P.E.M., S.A.Y., M.G., A.J., L.S.B., S.L., and R.A.S.; resources, G.A.Y. and R.A.S.; software, G.A.Y.; statistical analysis, G.A.Y.; interpretation of results, all authors; writing—original draft preparation, G.A.Y., P.B.J., A.M.M., U.B., S.P.E.M. and R.A.S.; writing—review and editing, all authors. All authors contributed important intellectual content and have read and agreed to the final version of the manuscript.

## SUPPLEMENTARY MATERIAL

The supplementary material for this article can be found at XXX.

